# Multivariate Profiling of Physical Resilience in Older Adults After Total Knee Replacement Surgery

**DOI:** 10.1101/2024.10.03.24314863

**Authors:** Qian-Li Xue, Thomas Laskow, Mallak K. Alzahrani, Ravi Varadhan, Jeremy D. Walston, Jennifer A. Schrack, Anne B. Newman, Frederick Sieber, Karen Bandeen-Roche

## Abstract

**IMPORTANCE:** As individuals age, they often face a variety of health challenges. Physical resilience indicates how well a person can cope with and recover from physical challenges, which is crucial for maintaining independence and quality of life in older age.

**OBJECTIVE:** To develop a multivariate phenotype of physical resilience based on individual recovery dynamics before and after a clinical stressor.

**DESIGN, SETTING, AND PARTICIPANTS:** This observational study included 112 individuals aged 60 and older who underwent elective total knee replacement for degenerative joint disease between December 2, 2019, and January 4, 2023. Physical function was assessed before surgery and at 1, 6, and 12 months post-surgery to characterize resilience trajectories.

**EXPOSURE:** Elective total knee replacement surgery for degenerative joint disease.

**MAIN OUTCOMES AND MEASURES:** A multivariate resilience phenotype was derived from physical function trajectories assessed using the Short Physical Performance Battery, the Pittsburgh Fatigability Scale-Physical Subscale, the KOOS Quality of Life, and the SF36-Physical Component Score. This phenotype was validated against surrogate markers (i.e., frailty, self-reported health) and determinants (e.g., the Charlson Comorbidity Index) of recovery potential (aka resilience capacity).

**RESULTS:** The study identified distinct resilience profiles across four measures: 4 profiles for the Short Physical Performance Battery and the KOOS Quality of Life, 3 each for the Pittsburgh Fatigability Scale-Physical Subscale and the SF36-Physical Component Score, showing varied baseline levels and/or change rates over 12 months. By combining and analyzing resilience profiles across measures, two distinct groups emerged: 35.7% classified as non-resilient and 64.3% as resilient. The non-resilient group had a higher prevalence of frailty (35.0% vs. 9.7%, p<0.01), poor or fair self-reported health (45.0% vs. 5.6%, p<0.01), and a moderate/severe comorbidity burden (Charlson Comorbidity Index >2; 27.5% vs. 11.1%, p=0.06).

**CONCLUSIONS AND RELEVANCE:** The distinct recovery trajectories observed after the surgery indicated varying resilience levels that were not fully explained by baseline status. This research underscores the importance of resilience in surgical recovery and could pave the way for better patient care by focusing on individual resilience capacities and shifting the focus from managing health conditions to promoting recovery and overall well-being.

**Key Points:** *Question:* Can recovery trajectories of physical function following total knee replacement surgery serve as indicators of resilience to physical stressors?

*Findings:* An observational study of adults aged 60+ undergoing elective total knee replacement surgery found distinct 12-month recovery paths, with 35.7% classified as non-resilient and 64.3% as resilient, independent of pre-surgery health or fitness.

*Meaning:* This finding suggests that resilience is measurable and may require dynamic testing, rather than just relying on baseline health, to assess recovery potential.

## Introduction

Resilience is an individual’s ability to adapt, recover, and return to equilibrium at molecular, cellular, system, organ, or organism level after experiencing a significant adversity or stress^1^ This paper focused on physical stressors, and therefore, “physical” resilience.^2^ We further posit that resilience differs from robustness (or resistance) in that the former refers to “bouncing back” following a stressor to retain “essential identity and function”,^3^ whereas the latter pertains to the ability to withstand stressors. For example, the rate and degree of recovery from an invasive surgery illustrates resilience, whereas humoral immunity to avoid symptomatic viral/bacterial infections exemplifies robustness. This distinction helps focus resilience on the characterization of response dynamics following the stressor.

Different approaches have been proposed to characterize physical resilience phenotypes, which can be broadly divided into two main types. The first type views resilience as a state that can be ascertained cross-sectionally at a specific point in time, using instruments such as the Physical Resilience Scale.^2^ The second type defines physical resilience as a dynamical entity based on the change observed before and after a stressor, necessitating an assessment prior to the stressor. We further divide the second type into two subtypes. The first subtype quantifies resilience by assessing either the absolute or percentage change from the pre-stressor baseline, within a designated post-stressor time window that holds clinical importance.^4^ This approach is suitable for examining the immediate dynamics of stress-response. For example, monitoring the initial few weeks following an organ transplant is critical to assessing the risk of acute rejection and longer-term survival and making an accurate prognosis. The second subtype focuses on risk profiling based on individual recovery trajectories, emphasizing longer term rather than immediate dynamics. This approach includes methods that generate either quantitative summaries of resilience (e.g., recovery differential)^5^ or qualitative resilience profiles (e.g., high vs. low resilience)^6^.

Choosing among the different approaches depends on the context in which they are used and the study design and data availability. The cross-sectional post-stressor ascertainment approach is suitable for situations involving an unpredictable stressor, such as a hip fracture, where assessing pre-stressor status is usually infeasible and self-recall may be unreliable.

Conversely, the trajectory approach is more appropriate for predictable stressors such as elective surgery, where it is possible to evaluate pre-stressor status. In terms of trajectory summary, the quantitative approach offers granularity, precision, and statistical power but requires advanced modeling techniques and can be complex to interpret in clinical settings. On the other hand, qualitative profiles, although easier to communicate and supportive of clinical decision-making, may involve complex computing algorithms and risk misclassification and reduced statistical power.

This study used data from one of the three substudies of the Study of Physical Resilience and Aging (SPRING)--RESilience in TOtal knee REplacement (RESTORE).^7^ We aimed to develop a multivariate phenotype of physical resilience for adults 60 and older undergoing elective total knee replacement (TKR) surgery for degenerative joint disease, based on their individual recovery dynamics before and after the surgical stressor. The primary utility of this phenotype is to provide a study outcome for validating measures of pre-stressor physical resilience capacity and its determinants (Figure 1). Additionally, it could serve as a prognostic tool for identifying vulnerable patients who may require special attention and interventions post-surgery in order to minimize adverse outcomes.

**Figure 1.**
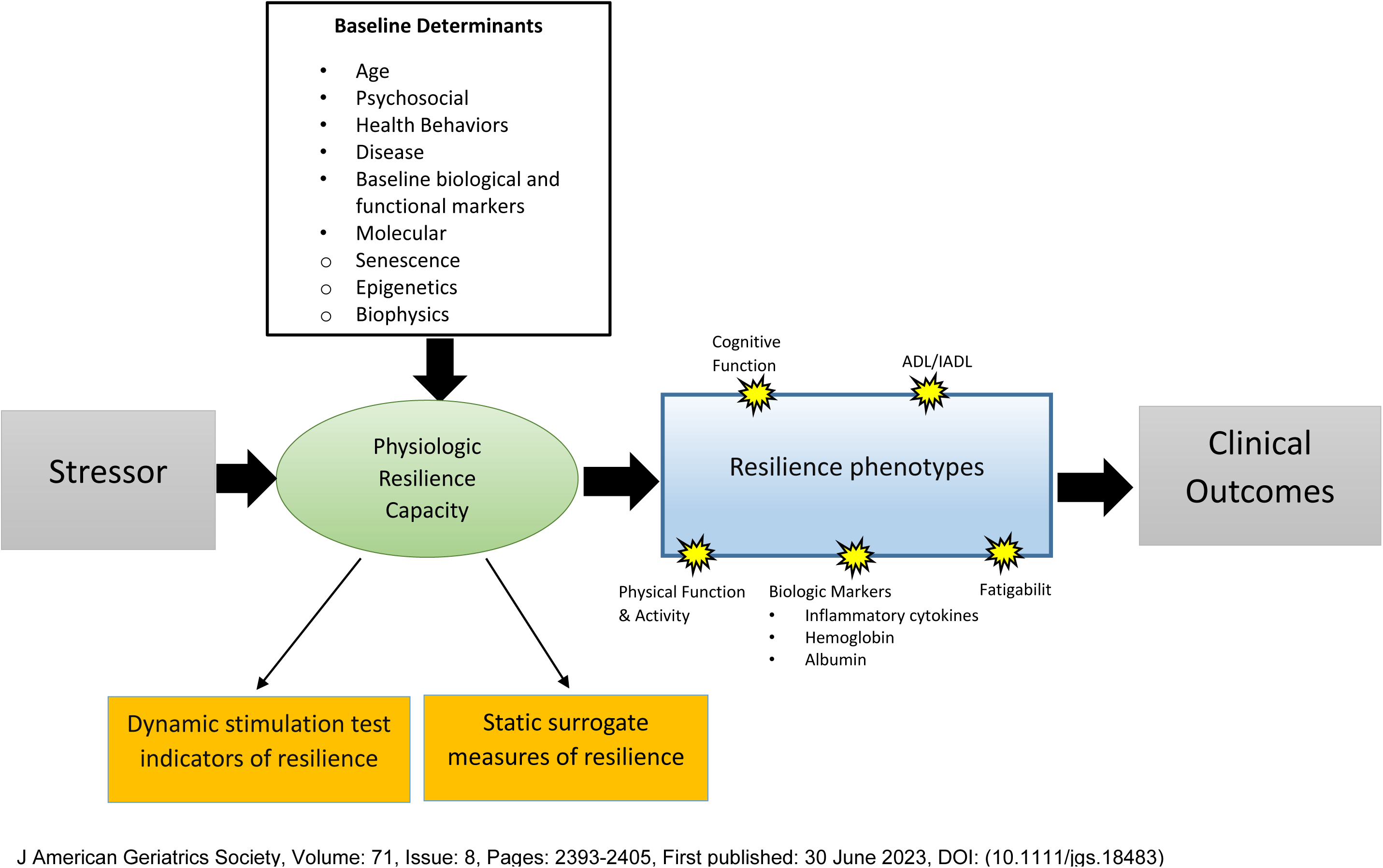
Conceptual Framework for Physical Resilience.

## Methods

### Study Population

SPRING was an observational study aimed at developing a framework to identify clinically relevant signatures of resilience in older adults facing physical stressors.^7^ Within SPRING, the RESilience in TOtal knee REplacement (RESTORE) substudy focused on characterizing older adults undergoing elective TKR surgery (eMethods 1). Extensive measurements were collected on a total of 112 older adults aged 60 years or older before, during, and after the surgical procedure to understand the impact of this stressor. This study was observational and did not influence surgical decisions.

### Clinical Assessments

Two baseline visits and follow-ups at 1, 6 & 12 months were used to assess resilience and related measures (eFigure 1). The resilience phenotype was derived from four measures the RESTORE team deemed particularly relevant for TKR resilience: the Short Physical Performance Battery (SPPB,^8^ the Pittsburgh Fatigability Scale – Physical Subscale (PFS),^9^ the Knee Injury and Osteoarthritis Outcome Score Quality of Life subscale (KOOS-QOL),^10^ and the physical component summary (PCS) of the SF-36 questionnaire.^11^ The SPPB assesses lower extremity function in older adults by evaluating their performance in three tasks simulating daily activities: balancing, standing from a chair, and walking, yielding a score from 0 to 12 with higher scores indicating better physical performance. The PFS is a self-administered 10-item questionnaire assessing perceived physical and mental fatigability related to fixed-intensity and duration activities, scored on a scale of 0 to 50 (reversed here, with higher scores indicating less fatigue). The KOOS-QOL measures the impact of knee injury or osteoarthritis on quality of life, transformed to a 0–100 scale with higher scores indicating better knee-related quality of life. The PCS of the SF-36 provides an overall view of perceived physical functioning based on a weighted sum of eight subscale scores and standardized to have a mean of 50 and a standard deviation of 10, with higher scores indicating better physical health.

According to the SPRING conceptual framework (Figure 1),^7^ surrogates and determinants of physical resilience were used to assess the convergent validity of the resilience phenotype. Frailty (surrogate) was assessed using the physical frailty phenotype,^12^ based on five criteria including unintentional weight loss (10 pounds or more in the past year), weakness (measured by reduced grip strength), poor endurance and energy (self-reported exhaustion or low energy levels), slowness (slow walking speed over 4 meters), and low physical activity (based on self-reported activity). Frailty and pre-frailty were determined by the presence of three or more, and one or two of these criteria, respectively. Self-reported overall health (surrogate) based on self-perception was assessed by self-report on a 5-point scale excellent, very good, good, fair, and poor. Disease burden (determinant) was measured by the Charlson Comorbidity Index (CCI). Comorbidity severity was graded as none (CCI=0), mild (CCI=1-2), moderate (CCI=3-4), and severe (CCI ≥5).^13^

### Statistical Analyses

We summarized the baseline characteristics for the study sample and compared them based on the overall resilience status using two-sample t-tests with unequal variances for continuous factors and Fisher’s exact tests for categorical factors. To develop resilience phenotypes using data from pre-surgery baseline and follow-ups at 1-, 6-, and 12-months post-surgery, we employed a two-stage latent variable model: a first-stage latent profile analysis (LPA),^14,15^ followed by a second-stage latent class analysis (LCA).^14^ The LPA was applied separately to each of the four phenotypic measures to capture distinct temporal trajectory profiles. In the second stage, the LCA aggregated these trajectory profiles across measures to derive a summary phenotype, distinguishing resilience from non-resilience (eMethods 2). Next, we assessed the convergent validity of the resilience phenotypes by analyzing their associations with the surrogates and determinant of resilience capacity identified above. MPLUS (version 8.10) was used to fit the latent variable model, and other analyses used Stata/SE 18.0.

## Results

Of the 112 participants in this study, the mean age was 70 years; 38% were Black and 59% were White. The group was predominantly female (66%) and had more than a high school education (62.5%), and nearly half (49%) were married. The mean BMI was 32. Regarding health status, 19% and 58% were frail and pre-frail, respectively; 38% reported excellent or very good health, and 22% reported fair or poor health. Additionally, 27% and 17% had mild and moderate/severe comorbidity burden based on CCI (Table 1).

**Table 1.** Baseline characteristics of study sample.

| **Characteristic** | **All**<br>**(N=112)** | **Non-Resilient**<br>**(n=40)** | **Resilient**<br>**(n=72)** | **P Value<sup>a</sup>** |
| --- | --- | --- | --- | --- |
| Age, mean (SD), year | 69.5 (6.9) | 69.4 (6.6) | 69.6 (7.1) | 0.906 |
| Sex, n (%) |  |  |  | 0.513 |
| Female | 74 (66) | 28 (38) | 46 (62) |  |
| Male | 38 (34) | 12 (32) | 26 (68) |  |
| Race, n (%) |  |  |  | 0.008 |
| White | 66 (59) | 16 (24) | 50 (76) |  |
| Black | 42 (38) | 23 (55) | 19 (45) |  |
| Other | 4 (4) | 1 (25) | 3 (75) |  |
| Education (grades completed), n (%) |  |  |  | 0.704 |
| <12 | 8 (7) | 3 (38) | 5 (63) |  |
| 12 | 34 (30) | 14 (41) | 20 (59) |  |
| >12 | 70 (63) | 23 (33) | 47 (67) |  |
| Family Income (US dollar), n (%) |  |  |  | 0.093 |
| <\$25K | 9 (8) | 5 (56) | 4 (44) | |
| ≥\$25K, <50K | 13 (12) | 4 (31) | 9 (69) | |
| ≥50K, <100K | 22 (20) | 11 (50) | 11 (50) |  |
| ≥100K | 39 (35) | 8 (21) | 31 (80) |  |
| Unknown | 29 (26) | 12 (41) | 17 (59) |  |
| Marital status, n (%) |  |  |  | 0.540 |
| Married | 55 (49) | 17 (31) | 38 (69) |  |
| Not married | 33 (30) | 14 (42) | 19 (58) |  |
| Widowed | 24 (21) | 9 (38) | 15 (63) |  |
| Body Mass Index: mean (SD) | 32.1 (5) | 33.5 (5) | 31.1 (6) | 0.030 |
| Charlson Comorbidity Index, n (%) |  |  |  | 0.064 |
| None | 63 (56) | 18 (29) | 45 (71) |  |
| Mild | 30 (27) | 11 (37) | 19 (63) |  |
| Moderate/Severe | 19 (17) | 11 (58) | 8 (42) |  |
| Physical Frailty, n (%) |  |  |  | 0.004 |
| Robust | 26 (23) | 6 (23) | 20 (77) |  |
| Pre-frail | 65 (58) | 20 (31) | 45 (69) |  |
| Frail | 21 (19) | 14 (67) | 7 (33) |  |
| Self-Reported Health, n (%) |  |  |  | <0.001 |
| Excellent/Very Good | 42 (38) | 6 (14) | 36 (86) |  |
| Good | 48 (43) | 16 (33) | 32 (67) |  |
| Fair/Poor | 22 (20) | 18 (82) | 4 (18) |  |

<sup>a</sup>Two-sample t-test with unequal variances for continuous variables and Pearson chi-square test for categorical variables

For all four phenotypic measures except KOOS-QOL, the majority of recovery occurred between one and six months post-surgery, while KOOS-QOL showed continued improvement through 12 months (Figure 2). The latent profile model identified four trajectory classes based on SPPB scores at baseline and during follow-ups (Figure 3A). Two classes represented the lowest (9.8%) and highest (35.7%) mean SPPB scores at baseline, both improving over time. The other two classes had similar intermediate baseline scores but showed differing trends over time: one with a stable SPPB trajectory (17.9%) and the other with an improving trajectory (36.6%). We identified three trajectory classes for PFS, primarily differentiated by varying levels of baseline physical fatigability (Figure 3B): low (23.2%), medium (63.4%), and high (13.4%). All classes showed improvements from one month to six months post-surgery. The analysis of KOOS-QOL scores over time identified four trajectory classes, all showing improvement (Figure 3C). The group with the lowest baseline mean score saw a moderate increase from baseline to 1 month, then plateaued (4.5%). The two classes with medium baseline scores exhibited steady growth until six months; one continued to rise (17.0%), while the other stabilized (22.3%). The largest group, with the highest baseline mean score, demonstrated consistent improvement throughout the 12 months (56.3%). Three trajectory classes for PCS were identified, each improving over time (Figure 3D). Two classes, one with the lowest baseline mean score (22.3%) and the other with the highest (34.8%), showed similar trends, with improvements noted between 1 and 6 months. The third class, featuring a similar baseline mean score as the lowest baseline class, demonstrated steady improvement from baseline to six months, accounting for 42.9% of the sample (eTable 1). The sensitivity analysis that treated the phenotypic measures as ordinal latent class indicators showed similar patterns (eFigure 2).

**Figure 2.**
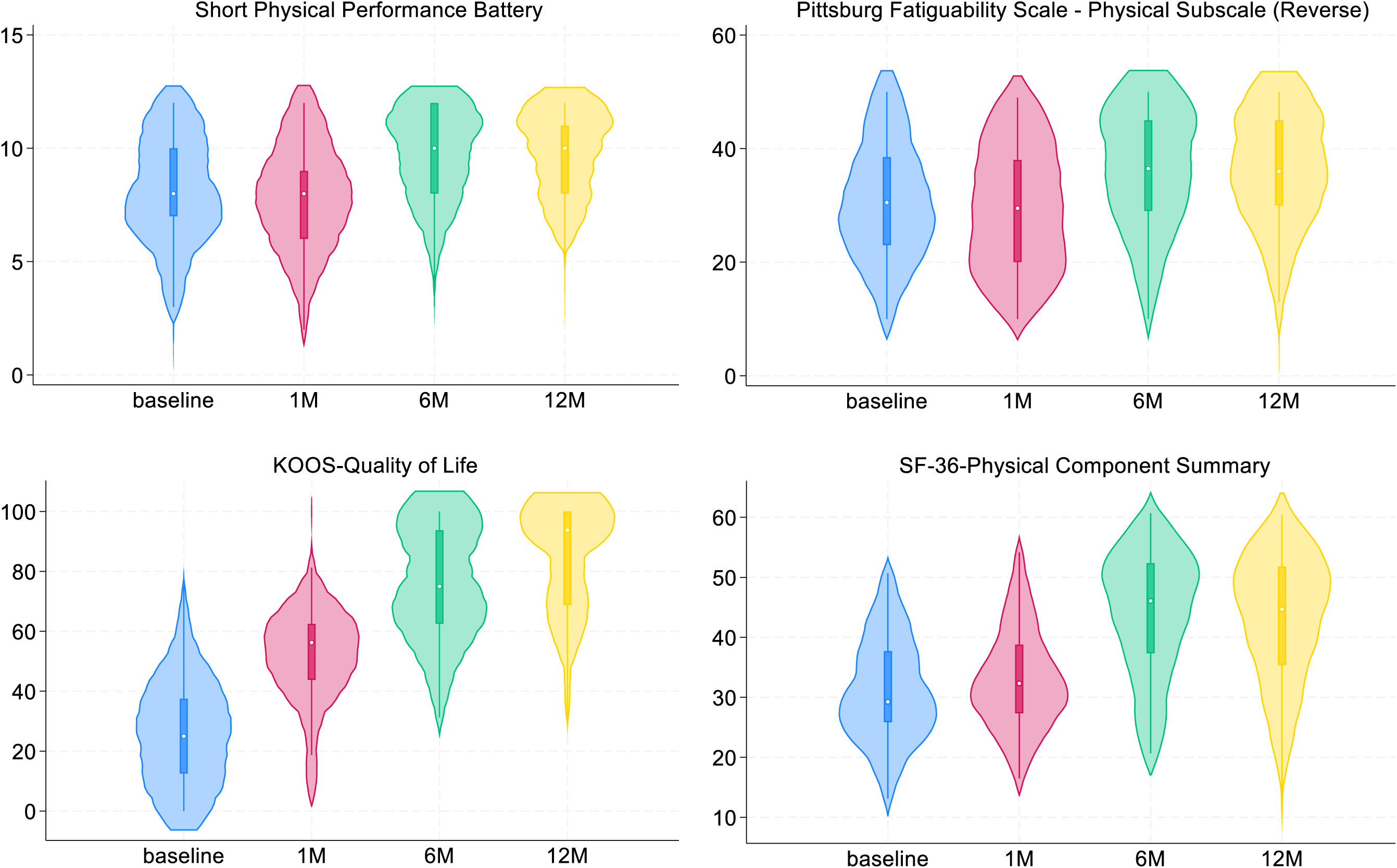
Marginal distribution of resilience phenotype measures by time in month.

**Figure 3.**
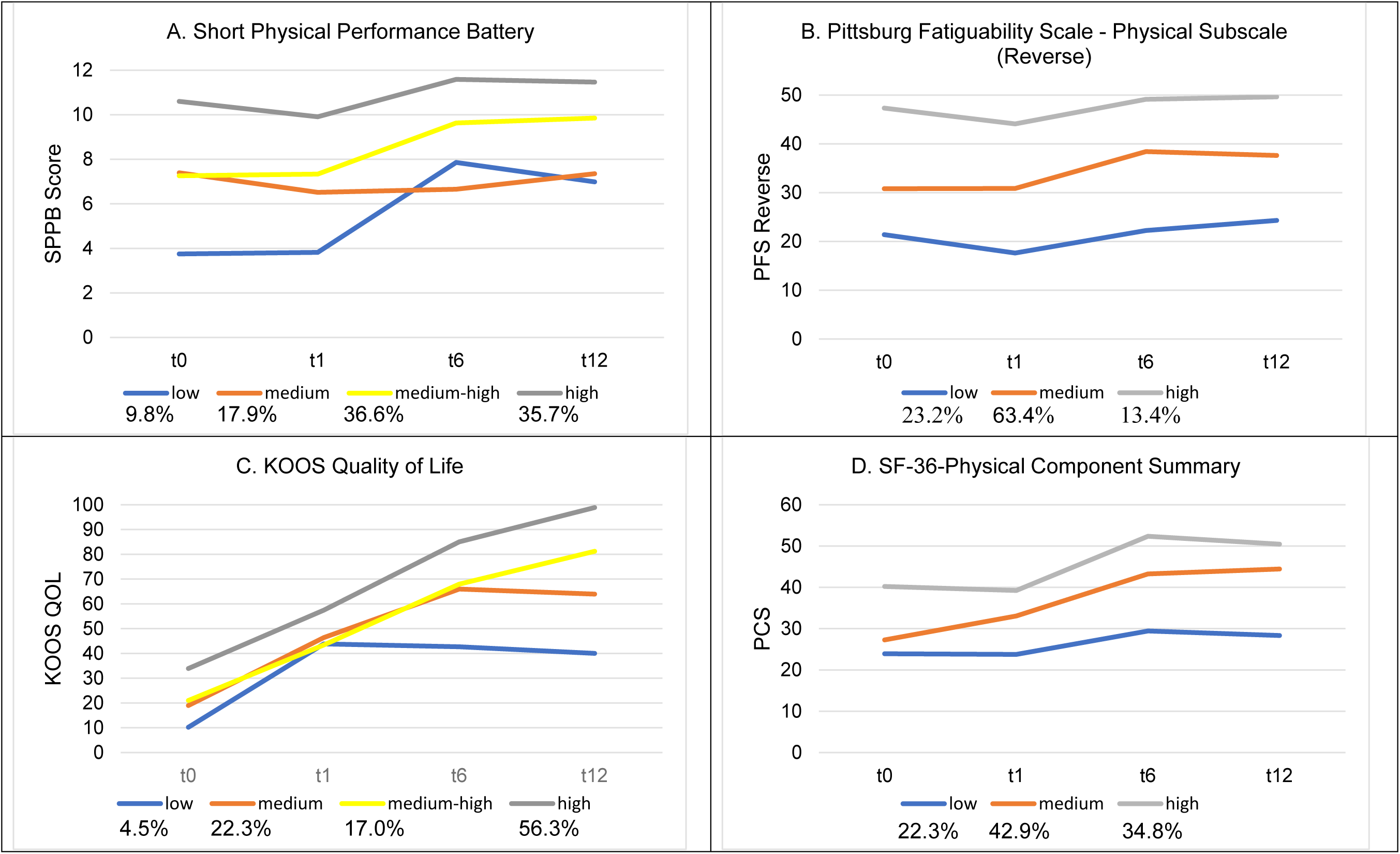
Patterns of phenotypic trajectories derived from latent profile analysis using continuous latent class indicators.

Latent class analysis of the four phenotypic measures identified two trajectory summary classes characterized by high (64.3%) and low (35.7%) resilience. Figure 4 compares phenotypic trajectory profile prevalence for resilient versus non-resilient individuals. There is one panel per measure: Each shows the percentages for each trajectory profile, progressing from the least favorable on the left to the most favorable on the right, comparing the resilient and non-resilient summary classes. Across all measures, individuals in the resilient group demonstrated more favorable trajectory patterns compared to those in the non-resilient group.

**Figure 4.**
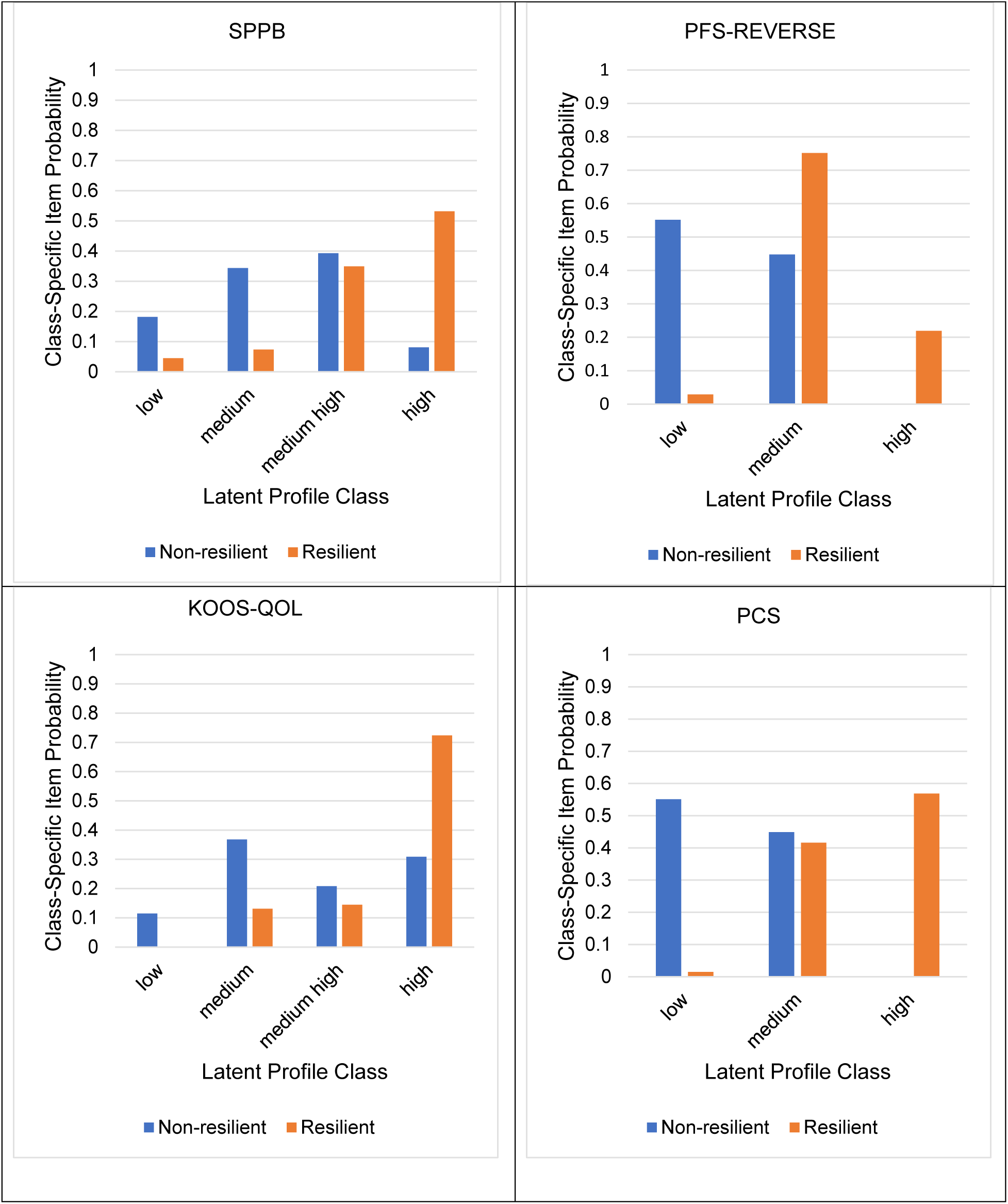
Trajectory profile composition by measure: progression from least to most favorable outcomes, comparing resilient and non-resilient classes.

Frailty was associated with low resilience, with 67%, 31%, and 23% frail, prefrail, and robust of individuals exhibiting a low resilience trajectory (p=0.004). Conversely, better self-reported health was associated with greater resilience, with 86%, and 67% of those in excellent/very good, and good health classified as having high resilience, compared to 18% of those in fair/poor health (p<0.001). These associations remained significant after adjusting for age, race, obesity, and disease burden. Specifically, frailty reduced the odds of high resilience by 79.3% (95% confidence interval (CI), 5.6-95.5%; p=0.042) compared to being robust, while excellent/very good health and good health increased the odds (Odds ratio (OR)=19.4, 95% CI, 4.0-92.8; p<0.001 and OR=10.1, 95% CI, 2.5-41.1; p=0.001), compared to poor/fair health. Greater disease burden was associated with lower resilience, with 58% of those with moderate/severe disease burden classified as having low resilience, compared to 37% and 29% for those with mild or no burden (p=0.064). After adjusting for age, race, and obesity, moderate/severe disease burden was associated with 72% lower odds of high resilience compared to no burden (95% CI, 12.5-91.0%; p=0.029).

## Discussion

In this observational study, we tracked the trajectories of physical function, beginning shortly before and continuing for twelve months after TKR. We identified distinct patterns, indicating varying resilience levels not fully explained by baseline status. The consistency across measures supports the concept of an overall physical resilience phenotype. Strong correlations between this resilience phenotype and both physical frailty and self-reported health, surrogates of resilience capacity, further validated its relevance to the intended measurement target.

In recent decades, studies have sought to better characterize recovery trajectories in individuals undergoing TKR. Some focused on the natural history and expected clinical course, while others examined early recovery to predict long-term outcomes. Our work aligns with the former approach but differs in two key ways: we broadened resilience measures beyond knee pain and dysfunction to include whole-body function, and we defined resilience at the whole-person level by analyzing trajectories across multiple functional measures. Our findings are consistent with others, showing that most improvement occurs within the first six months post-TKR. ^16–22^ Additionally, we identified subsets of older adults with similar pre-TKR function but diverging trajectories, highlighting variability in resilience. ^23–25^ The close alignment between overall resilience status and measure-specific resilience profiles supports the concept of an underlying resilience capacity influencing recovery across domains. This lays the foundation for future research into the biological underpinnings of physical resilience, particularly the integrity of the stress response system dynamics,^7^ with the goal of developing interventions to enhance resilience capacity in older adults before physical stressors occur.

Various approaches have been used to model between-person heterogeneity in resilience trajectories. They can be broadly classified into three categories: growth curve models,^26^ growth mixture models,^27^ and latent variable models. For example, random effects (aka multilevel) models assume a parametric form for trajectories, with parameters such as time slope and acceleration varying continuously across individuals, as seen in the “Expected Recovery Differential Approach.”^5^ Growth mixture models, in contrast, identify distinct subgroups with different trajectory patterns, sharing a common form but varying in baseline function and rates of change.^5^ We opted for LPA due to its flexibility in capturing trajectory patterns nonparametrically, followed by LCA to summarize patterns across measures. While this model provides qualitative insights into trajectory patterns, it doesn’t quantify recovery levels or intervention effectiveness. Instead, it is valuable for hypothesis testing concerning determinants of physical resilience and for validating the construct of pre-stressor resilience capacity.

Definitions of physical resilience vary in the literature. For example, Resnick et al. defined it as the ability to “overcome physical challenges encountered by a physically stressful event.”^2^ In contrast, Whitson et al. viewed it as “a characteristic at the whole person level which determines an individual’s ability to resist functional decline or recover physical health following a stressor.”^1^ The former focuses on the physical nature of the stressor, while the latter addresses physical health as the main domain of interest. We adopted the first perspective, recognizing that stressors, physical or not, exert their influence through a complex interplay of biological, psychosocial, environmental, and behavioral factors. Focusing solely to physical health may oversimplify this impact, overlooking roles of social, psychological, and recent health events in physical function.^28^ Defining physical resilience by the nature of the stressor allows consideration of multiple factors – physical, psychological, environmental, and societal – when designing interventions to improve outcomes for older adults with low resilience or promote resilience against physical challenges.

In selecting measures of physical resilience, several key criteria were considered to ensure both scientific robustness and clinical relevance. Physical function measures were prioritized due to their proven importance in maintaining independence and quality of life in older adults.^29–31^ Sensitivity to stress-related changes, including recovery potential, was also crucial.^32^ A balance was sought between general and condition-specific measures. In our study, we used both broad indicators of overall health (e.g., PCS) and knee-specific metrics (e.g., KOOS-QOL) for a comprehensive view of resilience beyond just knee mobility.^33^ To minimize confounding, we chose measures less influenced by lifestyle factors (e.g., physical activity levels) unrelated to health.^34^ Specificity was key, especially for stressor-specific resilience, as reflected in the inclusion of KOOS-QOL and SPPB. The measures also needed to demonstrate variability at baseline and over time to capture meaningful differences, as seen in the diverging trajectories in Figure 3. Both self-reports and objective performance measures were included to capture a full understanding of physical capabilities.^35^ Discrepancies between these could offer valuable insights into cognitive impairments, social influences, or compensatory mechanisms. Lastly, practical aspects such as feasibility and cost-effectiveness were considered, especially for measures sensitive to short-term dynamics within six months post-stressor.^36^ The focus on short-term outcomes was balanced with the need to predict longer-term clinical endpoints. In summary, these criteria establish a robust framework for assessing physical resilience in both clinical and research settings.

The study has severe strengths. First, the comprehensive assessment of function before and after a common clinical stressor provides a rare glimpse into the dynamics of impact and recovery in a real-world scenario. Second, the use of latent variable models generated both measure-specific and aggregate resilience phenotypes while accounting for measurement error in the phenotypic measures. Third, these phenotypes provide necessary means to validate the construct of resilience capacity and its determinants – central goals of the SPRING study. However, the study also limitations. The limited sample size necessitated more restrictive assumptions in the LPA such as fixing the variance of SPPB and KOOS-QOL across latent classes,^37^ potentially biasing class assignment.^38^ Nonetheless, the observed individual-specific trajectories aligned reasonably with classification ( see eFigures 3-6). Additionally, the low prevalence of certain classes, such as the 4.5% in the “low” resilience class for KOOS-QOL, may be unreliable. Sensitivity testing by merging this class with the adjacent “medium” resilience class reclassified four subjects as non-resilient but did not alter the status of those initially classified as low resilience. Third, resilience phenotypes may be context-specific. While appropriate for the RESTORE study, these measures might not suit other settings with high complication rates, like bone marrow transplants This underscores the need to tailor resilience definitions for different contexts, though the fundamental principles concerning phenotypic measures and modeling approach remain applicable. Finally, the focus on physical function limits generalizability beyond physical health, but this was intentional, given the nature of the stressor and the minimal observed impact (data not shown). Additionally, SPRING’s framework considers psychological well-being a determinant, not an indicator, of physical resilience.

In summary, our study revealed distinct recovery paths, showcasing various levels of resilience, which were not solely determined by pre-surgery fitness or health. This suggests that pre-stressor resilience capacity, a hypothesized key driver of recovery, may not be adequately captured by pre-stressor static function measures. Instead, resilience capacity reflects the integrity of interconnected physiological systems governing the stress-response that is crucial to recovery.^7^ This research highlights the importance of resilience in surgical recovery and could lead to improved patient care by focusing on individual resilience capacities. Future research into the factors contributing to resilience capacity in older adults could transform healthcare by shifting from merely managing symptoms and prolonging life to promoting recovery and improving quality of life for older adults.

## Data Availability

Data
Data available: Yes
Data types: Deidentified participant data, data dictionary
How to access data: The datasets generated and/or analyzed during this study are not publicly available to protect participant confidentiality. However, they can be obtained from the corresponding author upon reasonable request.
When available: With publication
Supporting Documents
Document types: None
Additional Information
Who can access the data: Researchers whose proposed use of the data has been approved.
Types of analyses: Secondary analyses.
Mechanisms of data availability: Data will be made available without investigator support upon approval of a proposal and execution of a signed data access agreement.

## Acknowledgment

None.

## Author Contributions

Q-L. X.: conception and design, analysis and interpretation of data, drafting and revising the article, final approval of the version to be published; T.L., J.O., J.A.S, A.B.N.: interpretation of data, revising the article for important intellectual content, final approval of the version to be published; M.K.Z.: analysis and interpretation of data, final approval of the version to be published; R.V., J.D.W., F.S., K.B-R: conception and design, interpretation of data, revising the article for important intellectual content, final approval of the version to be published.

## Data Access, Responsibility, and Analysis

Dr. Qian-Li Xue had full access to all the data in the study and takes responsibility for the integrity of the data and the accuracy of the data analysis.

## Data Sharing Statement

**Data**

**Data available:** Yes

**Data types:** Deidentified participant data, data dictionary

**How to access data:** The datasets generated and/or analyzed during this study are not publicly available to protect participant confidentiality. However, they can be obtained from the corresponding author upon reasonable request.

**When available:** With publication

## Supporting Documents

**Document types: None**

**Additional Information**

**Who can access the data:** Researchers whose proposed use of the data has been approved.

**Types of analyses:** Secondary analyses.

**Mechanisms of data availability:** Data will be made available without investigator support upon approval of a proposal and execution of a signed data access agreement.

