## Supplemental materials for "Multivariate Profiling of Physical Resilience in Older Adults After Total Knee Replacement Surgery"

eMethods 1. RESTORE Study Design

eMethods 2. Modeling of Resilience Phenotype

eTable 1. Goodness-of-fit statistics and class prevalence estimates for resilience measures: comparison across latent classes derived from the latent profile model fitting, with final model estimates in bold.

eFigure 1. Study visits and timeline.

eFigure 2. Marginal distribution of resilience phenotype measures by time in month.

eFigure 3. Patterns of phenotypic trajectories derived from latent class analysis using categorical latent class indicators.

eFigure 4. Alignment between latent profile model trajectory classes (left) and observed subject-specific trajectories (right) for short physical performance battery (SPPB) scores.

eFigure 5. Alignment between latent profile model trajectory classes (left) and observed subject-specific trajectories (right) for Pittsburg Fatigability Scale (PFS) reversed physical subscale scores.

eFigure 6. Alignment between latent profile model trajectory classes (left) and observed subject-specific trajectories (right) for KOOL Quality of Life (KOOS- QOL) subscale scores.

eFigure 7. Alignment between latent profile model trajectory classes (left) and observed subject-specific trajectories (right) for SF-36 Physical Component Summary (PCS) scores.

eMethods 1. RESTORE Study Design

SPRING was an observational study aimed at developing a framework to identify clinically relevant signatures of resilience in older adults facing physical stressors. Within SPRING, the RESilience in TOtal knee REplacement (RESTORE) substudy focused on characterizing older adults undergoing elective knee replacement surgery. Extensive measurements were collected on a total of 112 older adults before, during, and after the surgical procedure to understand the impact of this stressor. This study was non-interventional and did not influence surgical decisions. Eligible for RESTORE participants were 60 years or older at recruitment, had scheduled total knee replacement surgery at the Johns Hopkins Bayview Medical Center or the University of Maryland Medical Center, were capable of walking without assistance, and able to give informed consent for the study. Exclusion criteria included recent hospitalization for worsening of congestive heart failure or chronic obstructive pulmonary disease, severe sensory impairments, active inflammatory or autoimmune diseases, glucocorticoid use, allergic reactions to adrenocorticotropic hormone, symptomatic claudication or symptomatic peripheral arterial disease, infections requiring IV antibiotics, non-English proficiency, and cognitive limitations affecting consent comprehension. Eligibility was assessed during the initial orthopedic surgery clinic visit for knee replacement evaluation by review of the surgical schedules, where the research team reviewed demographic and clinical data to identify potential study candidates. Those meeting initial criteria received a brief study overview and a one-page summary. If confusion or comprehension issues were noted, pre-screening stopped. Otherwise, interest was gauged, and willing participants received a phone screening to confirm eligibility. Eligible participants provided written consent at the baseline visit. The study consists of two baseline visits and follow-ups at 1, 6 & 12 months as illustrated in eFigure 1.

eMethods 2. Modeling of Resilience Phenotype

To develop resilience phenotypes using data from pre-surgery baseline and follow-ups at 1-, 6-, and 12-months post-surgery, we employed a hybrid latent variable model. This model incorporates a first-stage latent profile analysis (LPA),^1,2^ applied separately to each of the four phenotypic measures, and a second-stage latent class analysis (LCA).^1^ The LPA uses repeated measurements of each measure as continuous indicators of a categorical latent variable, capturing different temporal trajectories, or latent classes. These indicators are assumed to be independent and normally distributed within each latent class. In our model, class-specific variances of class indicators were allowed to vary for PCS and PFS but were common for SPPB and KOOS-QOL to address convergence issues. We determined the number of latent classes based on criteria of the Bayesian Information Criterion (BIC), entropy, the Lo-Mendell-Rubin Adjusted Likelihood Ratio Test, and the parametric Bootstrapped Likelihood Ratio Test, along with scientific interpretation.^3-7^

After the LPA stage, a categorical indicator for each phenotypic measure was created, representing the latent class with the highest posterior probability of membership based on the LPA results and observed data. The second-stage LCA then used these four indicators of class membership as categorical inputs to identify an overarching resilience phenotype, aggregating the trajectory patterns of the four measures. Given the small sample size and the number of latent class indicators, the number of latent classes for this second-stage LCA was deliberately limited a priori to no more than two classes to ensure model identifiability. Participants were assigned to the class with the highest posterior probability.

The hybrid model was fit using the maximum likelihood with robust standard errors. Missing data in the phenotypic measures were addressed using full-information maximum likelihood for model parameter estimation under the assumption of data missing at random. To avoid the risk of converging on local rather than global maxima, we initiated 1000 random sets of starting values in the initial stage and conducted 50 optimizations in the final stage of modeling fitting for both the LPA and LCA. We evaluated the model’s goodness-of-fit by visually comparing the measure-and-class-specific trajectory patterns from the LPA to the observed subject-specific trajectories to assess their representativeness, as well as standardized residuals of the LCA. Two sensitivity analyses were conducted. First, a class with low prevalence (4.5%) from the LPA of KOOS-QOL was merged with an adjacent class to assess the impact of a small class on measure-specific and overall resilience status. Second, previous studies have demonstrated through data simulation that non-normal errors in regression mixture models can introduce biases in both model selection and parameter estimates (Van Horn et al. 2012). In this second sensitivity analysis, we addressed the skewed distribution caused by a ceiling effect in the phenotypic measures (eFigure 2) by categorizing these measures into tertiles or quartiles. Subsequently, we used LCA with ordinal latent class indicators instead of LPA in the initial stage. The LCA alternative has proven to be superior to other methods in modeling data affected by ceiling effects.^8^

**eTable 1. Goodness-of-fit statistics and class prevalence estimates for resilience measures: comparison across latent classes derived from the latent profile model fitting, with final model estimates in bold.**

| **Measure** | **Model Estimate** | **1-Class Model** | **2-Class Model** | **3-Class Model** | **4-Class Model** | **5-Class Model** |
| --- | --- | --- | --- | --- | --- | --- |
| SPPB | Class prevalence | 100% | 50.9%, 49.1% | 28.6%, 37.5%, 33.9% | **9.8%, 17.9%, 36.6%, 35.7%** | 9.8%, 17.9%, 27.7%, 22.3%, 22.3% |
|  | AIC | 1746.3 | 1591.7 | 1556.5 | **1528.7** | 1510.1 |
|  | BIC | 1768.0 | 1627.0 | 1605.4 | **1591.2** | 1589.2 |
|  | Adjusted BIC | 1742.7 | 1586.0 | 1548.5 | **1518.5** | 1497.8 |
|  | Entropy | NA | 0.84 | 0.80 | **0.87** | 0.87 |
|  | LMR | NA | 157.9 (p<0.01) | 43.4 (p=0.071) | **36.3 (p=0.082)** | 27.4 (p=0.398) |
|  | Bootstrap LRT | NA | 164.6 (p<0.01) | 45.2 (p<0.01) | **37.8 (p<0.01)** | 28.5 (p<0.01) |
| PFS^a^ | Class Prevalence | 100% | 74.1%, 25.9% | **23.2%, 63.4%, 13.4%** | 20.5%, 51.8%, 16.1%, 11.6% | NA |
|  | AIC | 3160.3 | 2985.2 | **2910.3** | 2887.3 |  |
|  | BIC | 3182.1 | 3031.4 | **2981.0** | 2982.5 |  |
|  | Adjusted BIC | 3156.8 | 2977.7 | **2898.8** | 2871.8 |  |
|  | Entropy | NA | 0.94 | **0.90** | 0.87 |  |
|  | LMR | NA | 188.7 (p<0.01) | **90.8 (p=0.011)** | 40.1 (p=0.084) |  |
|  | Bootstrap LRT | NA | 193.1 (p<0.01) | **92.9 (p<0.01)** | 41.0 (p=0.03) |  |
| KOOS-QOL | Class Prevalence | 100% | 33.9%, 66.1% | 4.5%, 34.8%, 60.7% | **4.5%, 22.3%, 17.0%, 56.3%** | 4.5%, 15.2%, 11.6%, 12.5%, 56.3% |
|  | AIC | 3589.9 | 3507.8 | 3481.9 | **3437.5** | 3426.7 |
|  | BIC | 3611.7 | 3543.1 | 3530.8 | **3500.1** | 3502.8 |
|  | Adjusted BIC | 3586.4 | 3502.1 | 3473.9 | **3427.4** | 3414.3 |
|  | Entropy | NA | 0.80 | 0.88 | **0.91** | 0.90 |
|  | LMR | NA | 88.4 (p=0.025) | 36.0 (p=0.10) | **52.1 (p=0.044)** | 20.0 (p=0.356) |
|  | Bootstrap LRT | NA | 92.1 (p<0.01) | 36.0 (p<0.01) | **54.3 (p<0.01)** | 20.8 (p=0.013) |
| PCS^a^ | Class prevalence | 100% | 51.8%, 48.2% | **22.3%, 42.9%, 34.8%** | 22.3%, 44.6%, 18.8%, 14.3% | NA |
|  | AIC | 3045.7 | 2902.9 | **2849.2** | 2854.3 |  |
|  | BIC | 3067.4 | 2949.1 | **2919.9** | 2916.8 |  |
|  | Adjusted BIC | 3042.1 | 2895.4 | **2837.7** | 2844.2 |  |
|  | Entropy | NA | 0.79 | **0.87** | 0.83 |  |
|  | LMR | NA | 157.0 (p=0.010) | **70.1 (p<0.01)** | 17.7 (p=0.091) |  |
|  | Bootstrap LRT | NA | 160.7 (p<0.01) | **71.8 (p<0.01)** | 18.4 (p=0.020) |  |

SPPB: Short Physical Performance Battery; PFS: Pittsburg Fatiguability Scale – physical subscale; KOOS-QOL: KOOS Quality of Life subscale; PCS: SF-36 Physical Component Summary; AIC: Akaike Information Criterion; BIC: Bayesian Information Criterion; LMR: Lo-Mendell-Rubin Test; LRT: likelihood ratio test.

^a^ Class-specific variances of class indicators were allowed to vary for PCS and PFS but were uniform for SPPB and KOOS-QOL to address convergence issues

**eFigure 1. Study visits and timeline.**

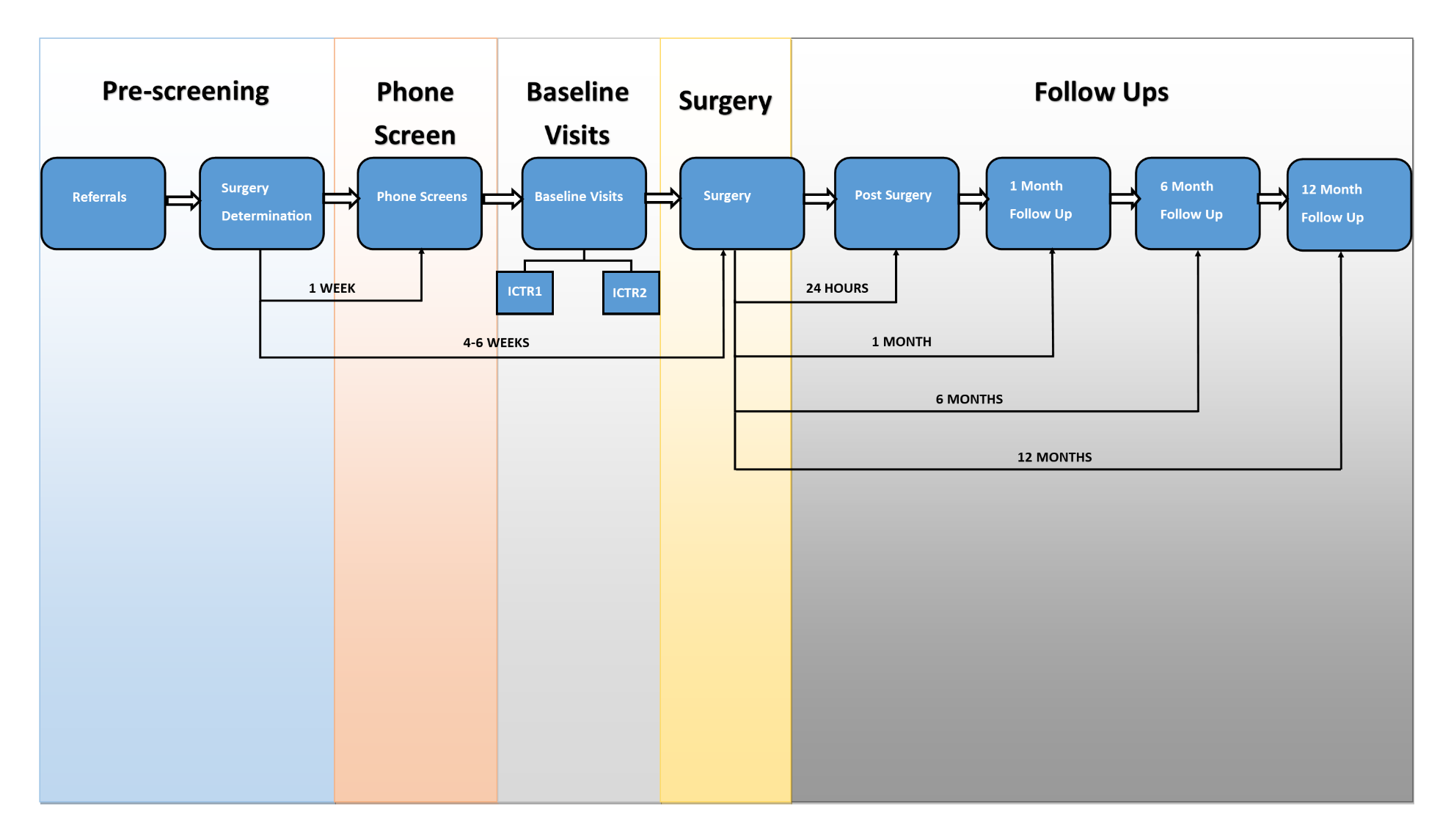

**eFigure 2. Patterns of phenotypic trajectories derived from latent class analysis using categorical latent class indicators.**

A. Short Physical Performance Battery

SPPB Score

|  | low | medium | medium high | high |
| --- | --- | --- | --- | --- |
| t0 | 4.281 | 7.057 | 7.002 | 10.4 |
| t1 | 3.5 | 6.299 | 6.9 | 10.017 |
| t6 | 9 | 6.4675 | 10.175 | 10.7395 |
| t12 | 7.523 | 7.917 | 10.0855 | 11 |

B. Pittsburg Fatiguability Physical Subscale (Reverse)

PFS Reverse Score

|  | low | medium | high |
| --- | --- | --- | --- |
| t0 | 20.5358 | 23.3483 | 42.066 |
| t1 | 16.5742 | 25.1197 | 41.6565 |
| t6 | 19.0487 | 37.442 | 43.8795 |
| t12 | 22.7218 | 36.871 | 43.033 |

5.4%	19.6%	17.9%	57.1%

C. KOOS Quality of Life

KOOS-QOL Score

|  | low | medium | medium high | high |
| --- | --- | --- | --- | --- |
| t0 | 18.9938 | 15.6629 | 11.5989 | 36.7725 |
| t1 | 46.0543 | 48.4323 | 42.5728 | 56.4808 |
| t6 | 51.9712 | 59.1466 | 71.3595 | 84.3096 |
| t12 | 54.2506 | 59.6104 | 91.4755 | 93.1542 |

D. SF-36-Physical Component Summary

PCS Score

|  | low | medium | high |
| --- | --- | --- | --- |
| t0 | 22.3818 | 30.1797 | 39.5475 |
| t1 | 22.5417 | 30.2923 | 42.549 |
| t6 | 25.0556 | 45.4597 | 47.43 |
| t12 | 25.5075 | 44.8535 | 47.43 |

**eFigure 3. Alignment between latent profile model trajectory classes (left) and observed subject-specific trajectories (right) for short physical performance battery (SPPB) scores.**

SPPB Score

|  | low | medium | medium-high | high |
| --- | --- | --- | --- | --- |
| t0 | 3.751 | 7.397 | 7.259 | 10.603 |
| t1 | 3.82 | 6.519 | 7.339 | 9.912 |
| t6 | 7.861 | 6.658 | 9.634 | 11.589 |
| t12 | 6.99 | 7.356 | 9.853 | 11.467 |

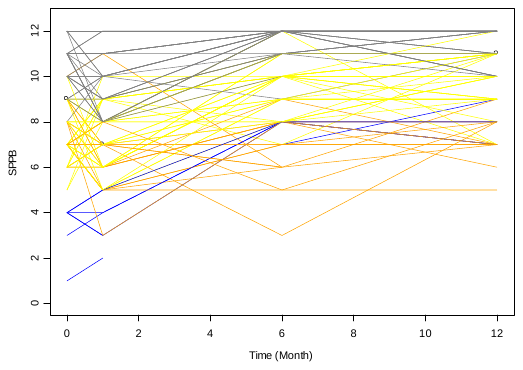

**eFigure 4. Alignment between latent profile model trajectory classes (left) and observed subject-specific trajectories (right) for Pittsburg Fatigability Scale (PFS) reversed physical subscale scores.**

PFS Reverse

|  | low | medium | high |
| --- | --- | --- | --- |
| t0 | 21.391 | 30.819 | 47.319 |
| t1 | 17.631 | 30.868 | 44.096 |
| t6 | 22.247 | 38.427 | 49.14 |
| t12 | 24.319 | 37.621 | 49.642 |

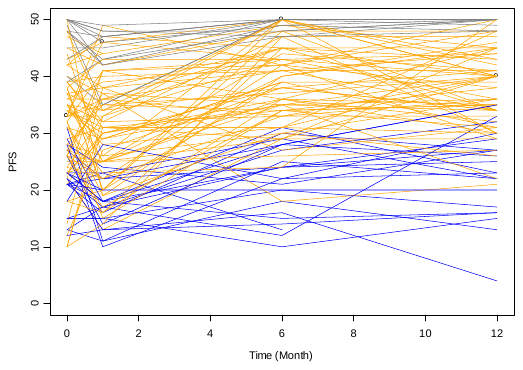

**eFigure 5. Alignment between latent profile model trajectory classes (left) and observed subject-specific trajectories (right) for KOOL Quality of Life (KOOS- QOL) subscale scores.**

KOOS QOL

|  | low | medium | medium-high | high |
| --- | --- | --- | --- | --- |
| t0 | 10.224 | 18.98 | 21.025 | 33.937 |
| t1 | 43.843 | 46.382 | 43.397 | 57.477 |
| t6 | 42.691 | 65.984 | 67.939 | 85.054 |
| t12 | 40.001 | 63.957 | 81.226 | 98.888 |

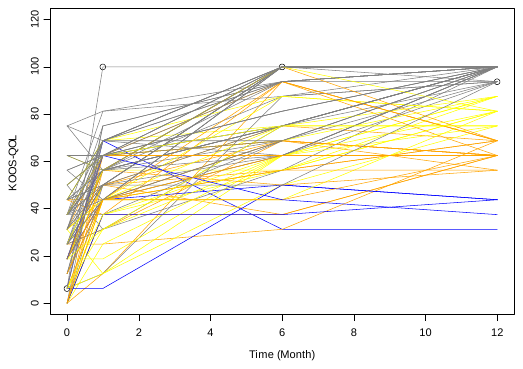

**eFigure 6. Alignment between latent profile model trajectory classes (left) and observed subject-specific trajectories (right) for SF-36 Physical Component Summary (PCS) scores.**

PCS

|  | low | medium | high |
| --- | --- | --- | --- |
| t0 | 23.931 | 27.273 | 40.228 |
| t1 | 23.756 | 33.061 | 39.239 |
| t6 | 29.424 | 43.259 | 52.365 |
| t12 | 28.336 | 44.438 | 50.448 |

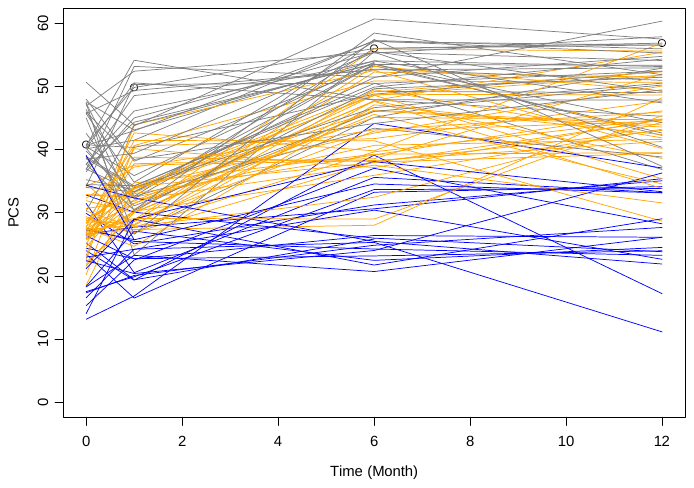
